# Erectile Dysfunction in Men with and without Type 2 Diabetes Mellitus at Livingstone University Teaching Hospital, Zambia: A cross-sectional study

**DOI:** 10.1101/2024.02.28.24303494

**Authors:** Lweendo Muchaili, Bislom C. Mweene, Benson M. Hamooya, Sepiso Kenias Masenga

## Abstract

**Background:** Erectile dysfunction (ED) is frequently undiagnosed in males with type 2 diabetes mellitus (T2DM), despite its high prevalence. Early detection of ED in T2DM is crucial for effective treatment and prevention of severe complications such as cardiovascular events. This study explores the prevalence of ED and its associated factors in males with and without T2DM at Livingstone University Teaching Hospital (LUTH), Zambia.

**Methodology:** We conducted a cross-sectional study at LUTH among 42 males aged ≥18 years (22 with and 20 without T2DM). T2DM diagnosis was confirmed through medical records, and erectile dysfunction was assessed using the International Index of Erectile Function (IIEF-5) questionnaire. Logistic regression identified factors associated with ED, with significance set at a p-value less than 0.05

**Results:** The overall prevalence of ED was 74% (31/42). Among T2DM participants, the prevalence of ED was 91% (20/22; 95% confidence interval (CI) 70.8-98.9) whereas among the non-diabetic participants, the prevalence was 55% (11/20; 95%CI 31.5-76.9). The majority [40% (8/20)] of the T2DM participants with ED had mild ED, 35% (7/20) had mild to moderate ED, 15% (3/20) had moderate ED and 10% (2/20) had severe ED. 54.5% (6/11)] of the non-T2DM participants with ED had mild ED, 36.4% (4/11) had mild to moderate ED, 9.1% (1/11) had moderate ED, and none had severe ED. In the multivariable analysis, employed individuals had lower odds of erectile dysfunction compared to the unemployed (OR 0.01, 95% CI 0.00 - 0.84, p = 0.041); while elevated plasma creatinine levels were associated with an increased risk of erectile dysfunction (OR 1.22, 95% CI 1.03 - 1.45, p = 0.021).

**Conclusion:** This study underscores a significant prevalence of ED, particularly heightened in T2DM participants; and significantly associated with plasma creatinine levels and employment status. The findings highlight the need for comprehensive assessment and management of ED in T2DM individuals. There is a need for further research with larger sample sizes to validate the findings and for a clearer understanding of associated factors and identification of effective targeted interventions.

## Introduction

Erectile dysfunction (ED) is defined as an inability to initiate and have a persistent erection firm enough to have satisfying sexual intercourse ^1^. In 1995 over 152 million people worldwide had ED and the number was projected to reach 322 million by the year 2025 with most new cases being in Africa, Asia, and South America ^2^. Some studies have reported that more than 50% of males with a diagnosis of ED have diabetes mellitus ^3^. Individuals with diabetes are 3 times more likely to have ED than unaffected individuals ^4^. . ED is defined as an inability to initiate and have a persistent erection firm enough to have satisfying sexual intercourse ^1^.

ED diagnosis has shown to be of value in screening for T2DM in presumably healthy males. T2DM males are 3 times more likely to develop ED than those without ED ^5^. ED diagnosis has also been recommended for coronary artery disease and related cardiovascular conditions in both diabetic and non-diabetic male patients ^6^. This is because some mechanisms that lead to ED such as hyperlipidemia and atherosclerosis are also associated with coronary artery disease and other cardiovascular diseases ^7,8^.

Studies have shown a strong correlation between uncontrolled T2DM and ED, thus ED may act as an indicator of poorly controlled diabetes and a warning sign for possible negative disease outcomes ^9^. ED in T2DM is as low as 35% in some populations and as high as 90% in others ^10–12^. ED in diabetes has a high prevalence in populations where there is poor hyperglycemic control and/or obesity and is often associated with hypertension, hyperlipidemia, autonomic neuropathy, and metabolic syndrome which are independent risk factors for cardiovascular disease^4,9^.

Previously, it has been shown that ED in T2DM is an early warning sign of impending major complications in T2DM such as cardiovascular diseases, coronary heart disease, and metabolic syndrome ^13–16^. Early diagnosis of ED in T2DM helps healthcare providers alter patient management. This may include the patient’s treatment regimen and the patient’s adjusting lifestyle. Controlling lifestyle has proved to be beneficial in improving erectile function and preventing major complications of T2DM ^4^.

Most underlying pathological mechanisms of ED and many cardiovascular diseases are the same, however, ED occurs earlier ^16^. The mean interval between the onset of ED and Coronary artery disease is about 24 months ^8,17^.

ED in T2DM in many African countries is high but diagnosis is not routinely done. Most cases of ED remain undiagnosed and its prevalence remains unknown ^18^. However, ED diagnosis is relatively simple and can be easily done using the International Index of Erectile Function (IIEF) questionnaire ^1^. The goal of this study was to determine the prevalence and associated factors of ED at Livingstone University Teaching Hospital (LUTH).

## Methods

### Study design and site

This was a cross-sectional study among adults with and without T2DM aged ≥18 years conducted at LUTH in the Livingstone district of Zambia between 1^st^ April 2023 and 30^th^ April 2023. LUTH is the regional referral health facility for Southern province. LUTH operates an outpatient department where out-patients access their medical services. The facility also operates a diabetic clinic where T2DM patients receive specialized medical services.

### Study population

The study comprised male individuals who were diabetic and non-diabetic and attending the outpatient department at LUTH.

### Eligibility criteria

#### Inclusion criteria

The study included all T2DM male adults and a control group of non-diabetic males matched for age. All participants were aged ≥18 years and sexually active. Only participants who agreed to sign the written consent were enrolled in the study.

#### Exclusion criteria

The study excluded clients who had a history of lower urinary tract surgery, past pelvic fracture, past spine injury or surgery, and those who were bedridden or critically ill.

#### Selection of participants and Sampling methods

For T2DM male participants, a census was conducted for males with T2DM (n=22) at LUTH. Due to the low number of males enrolled in the diabetes clinic, we offered to enroll all males with T2DM attending routine clinical visits at LUTH. Non-diabetic participants (n=20) were conveniently selected from the outpatient department and matched for age.

### Variables of interest

#### Primary outcome variable

The primary outcome variable was ED which was defined as the inability to initiate and have a persistent erection firm enough to have satisfying sexual intercourse ^19^. A participant either had ED or not according to their score on the IIEF - 5. The IIEF-5 score is the sum of the ordinal responses to the 5 items; the IIEF-5 score ranges from 5-25, with higher (22-25) scores denoting absence; and lower scores (5-21) denoting presence.

### Severity of erectile dysfunction

The severity of ED was graded as follows; 22-25, No erectile dysfunction; 17-21, Mild erectile dysfunction;12-16, Mild to moderate erectile dysfunction; 8-11, Moderate erectile dysfunction; 5-7, severe erectile dysfunction.

#### Independent variables

The independent variables were age, employment status, body mass index (BMI), diabetes status, hypertension status, hemoglobin, plasma creatinine, total cholesterol, urine protein, and urine ketones, lifestyle habits; smoking, alcohol consumption, frequency of eating vegetables, and frequency of physical exercise.

#### Definition of key variables

T2DM and hypertension diagnosis were based on past diagnosis including use of antihypertensive and diabetic drug history as contained in the medical records. BMI was calculated by dividing an individual’s weight in kilograms by the square of their height in meters. The formula is BMI = weight (kg) / height (m) ^2^. BMI was expressed in units of kg/m^2^. To determine the hemoglobin level of the participants, 4 ml of blood was collected in an EDTA container and analyzed on Sysmex XN 1000. Hemoglobin results were expressed in g/dL. For total cholesterol and plasma creatinine estimation, 4ml of blood in a plain container was collected and analyzed on Pentra C200. Creatinine was expressed in units of µmol/l, and total cholesterol in mg/dL. 15ml of urine was collected for urinalysis to determine the presence of ketones, glucose, proteins, and leukocytes using the 13-parameter urine multistix. These were qualitatively expressed as either negative or positive.

#### Data collection, tools, management, and storage

Interviewer-structured questionnaires were used to collect demographic and health information. The International Index for Erectile Function version 5 (IIEF-5) questionnaire was used to assess erectile function in the participants. The collected data were de-identified and stored entered into the Research Electronic Data Capture (REDCap) system. Only the Principal investigator and study assistants had access to the stored data.

#### Data analysis plan

STATA was used to analyze the data for this study. Descriptive statistics (frequency, medians, and proportions) were used for data description and for estimating the proportion of individuals with ED. The Wilcoxon rank sum test (to compare medians between two groups) and Kruskal-Wallis (to compare medians between more than two groups) were used to make inferences. A Chi-square test was used to compare proportions between outcome variables (ED) and categorical independent variables such as sex, education, employment status, and residential area. A logistic regression model (Univariable and multivariable) was used to examine factors associated with ED and to control for confounders. A p-value of less than 0.05 was considered statistically significant.

## RESULTS

### Basic characteristics of the study participants

The study comprised 42 male participants of whom 52.4% (22/42) had T2DM whereas 47.6% (20/42) were non-T2DM. The median age was 52.5 years [inter-quartile range (IQR) 39, 66]. The majority, 78.57% (33/42) of the participants were nonsmokers, never consumed alcohol 69.05% (29/42), and had no hypertension 61.9% (26/42), see Table 1.

**Table 1:**
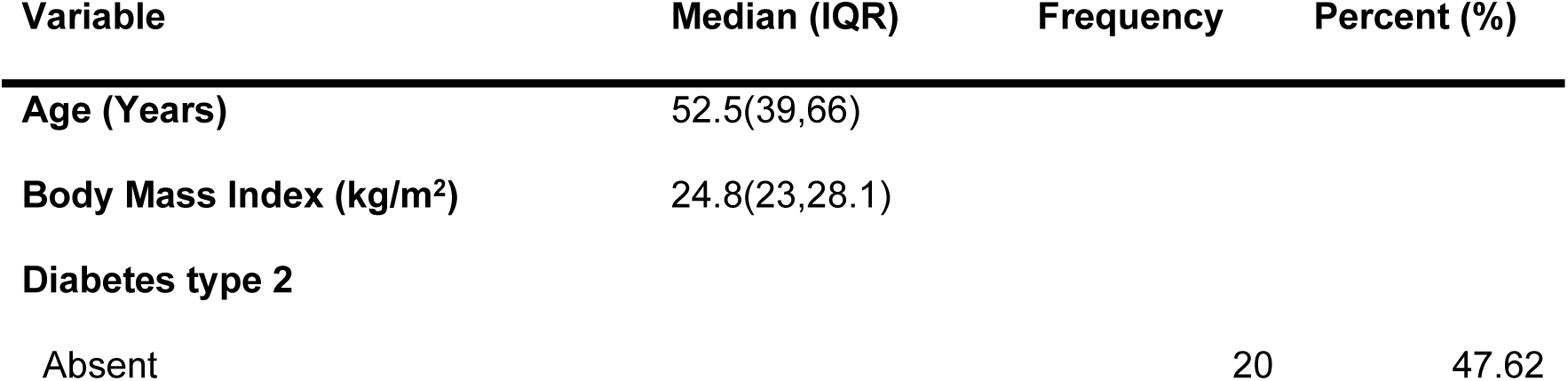
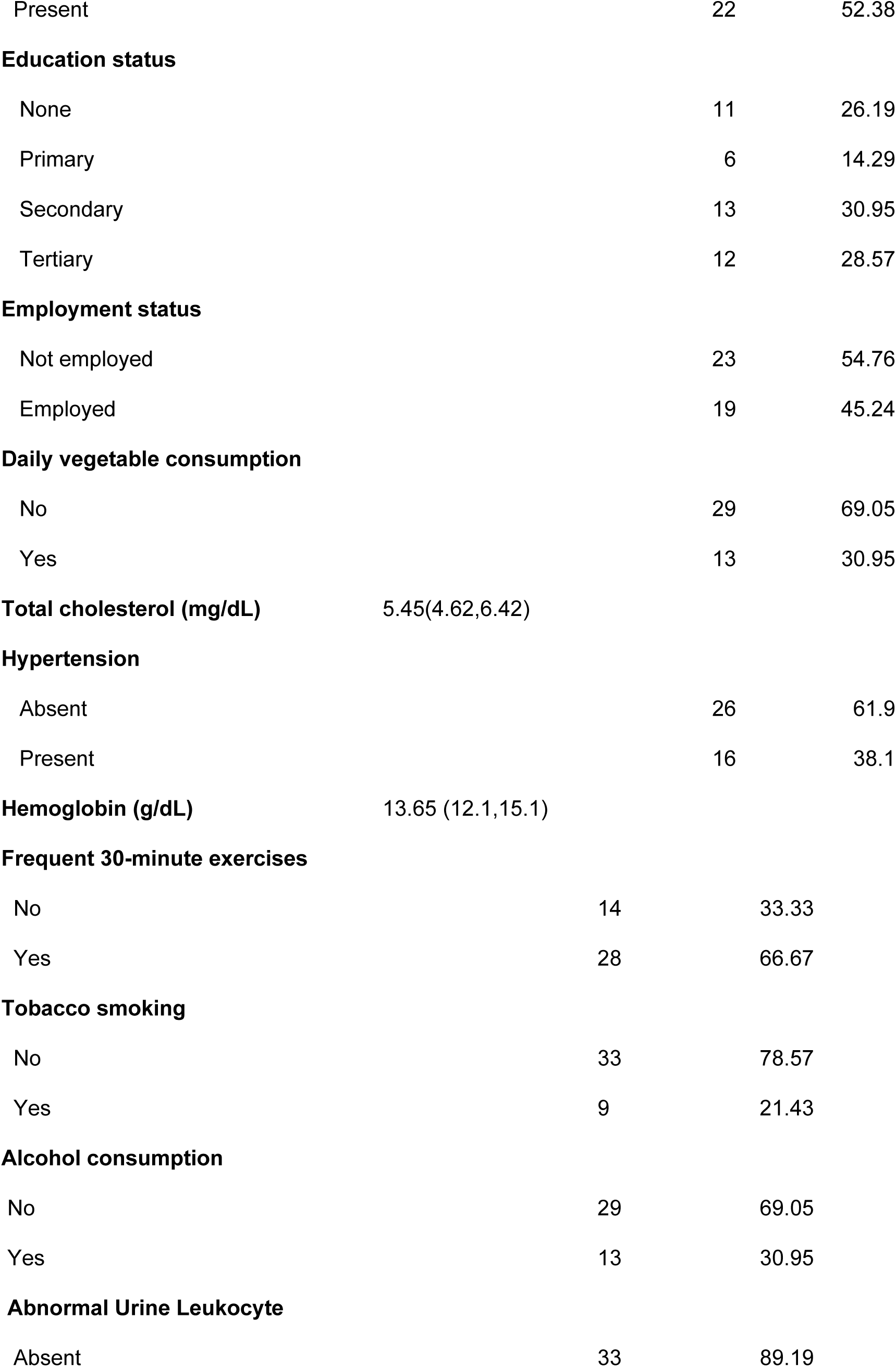
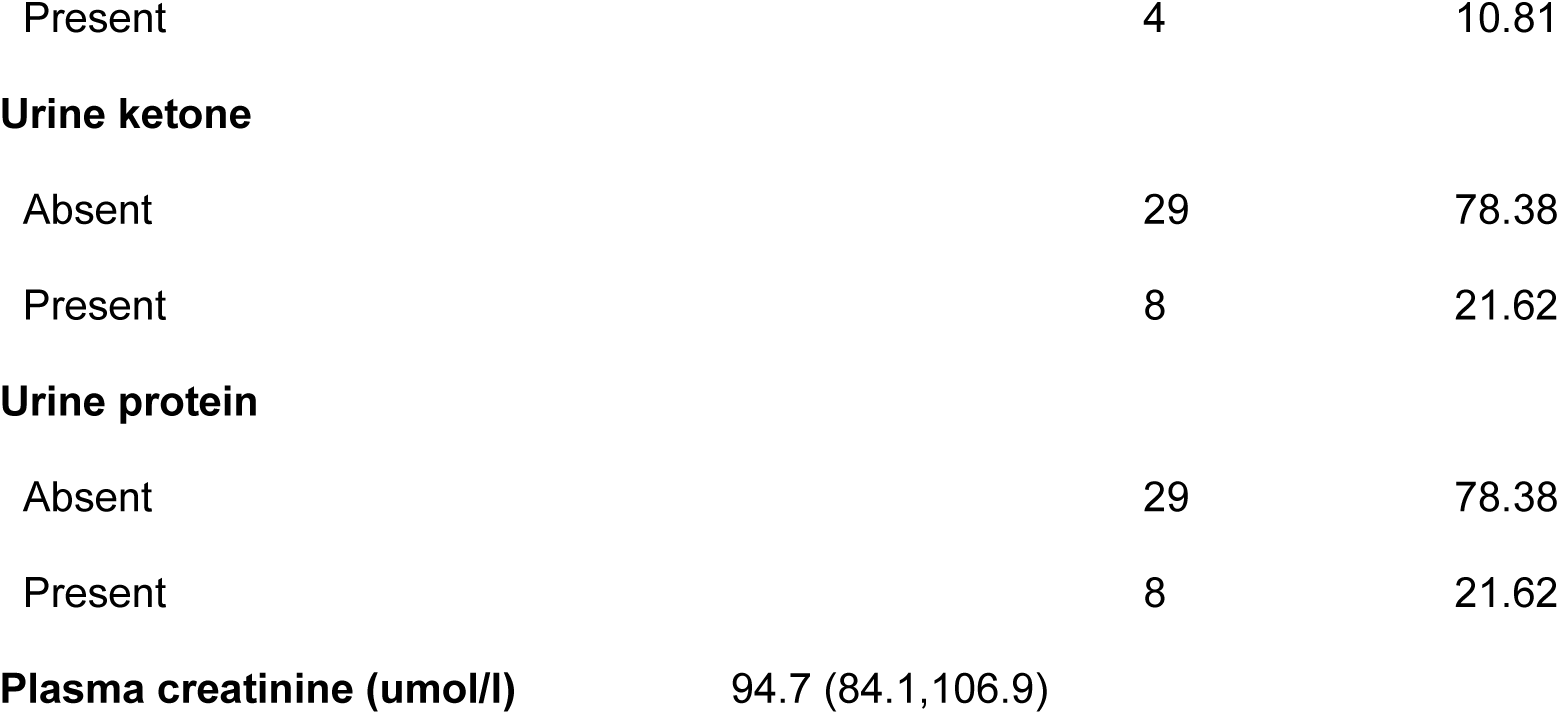
Demographic and clinical characteristics of the study participants.

#### Prevalence of Erectile dysfunction among the study participants

The overall prevalence of ED was 74% [31/42, (95% CI 0.57, 0.86)]. Among T2DM participants, the prevalence of ED was 91% [20/22, (95% CI 0.84, 1)] whereas among the non-T2DM participants, the prevalence was 55% [11/20, (95% CI 0.31, 0.76)].

#### Severity of ED among T2DM participants

Forty percent (8/20) of the T2DM male participants with ED had mild ED, 35% (7/20) had mild to moderate ED, 15% (3/20) had moderate ED and 10% (2/20) had severe ED. While 54.5% (6/11)] of the non-T2DM participants with ED had mild ED, 36.4% (4/11) had mild to moderate ED, 9.1% (1/11) had moderate ED, and none had severe ED. See Figure 1.

**Figure 1:**
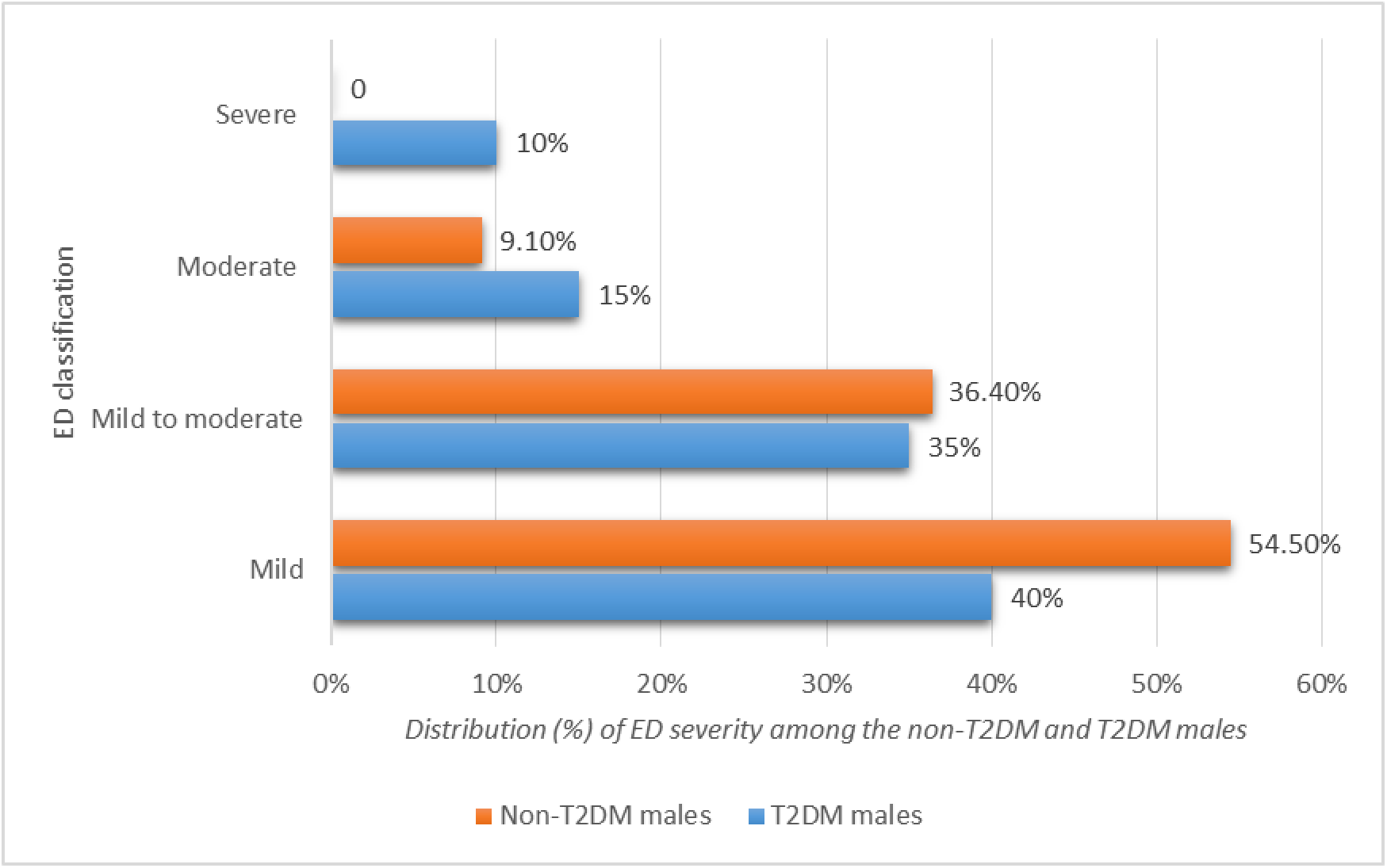
Severity of ED among participants with and without T2DM.

### Relationship between Erectile dysfunction and demographic and clinical factors

Table 2 shows factors associated with erectile dysfunction among the study participants. The study showed an association between age and ED, individuals with ED were significantly older than those without ED (median age 58 years vs. 39 years; p=0.002). There was also a significant association between employment status and ED, unemployed participants were more likely to have ED compared to their employed counterparts (83% vs. 70%, p = 0.005). Hypertension was also associated with ED, individuals with hypertension were more likely to have ED when compared to the non-hypertensive participants (93.75% vs. 61.5%, p=0.021).

**Table 2:**
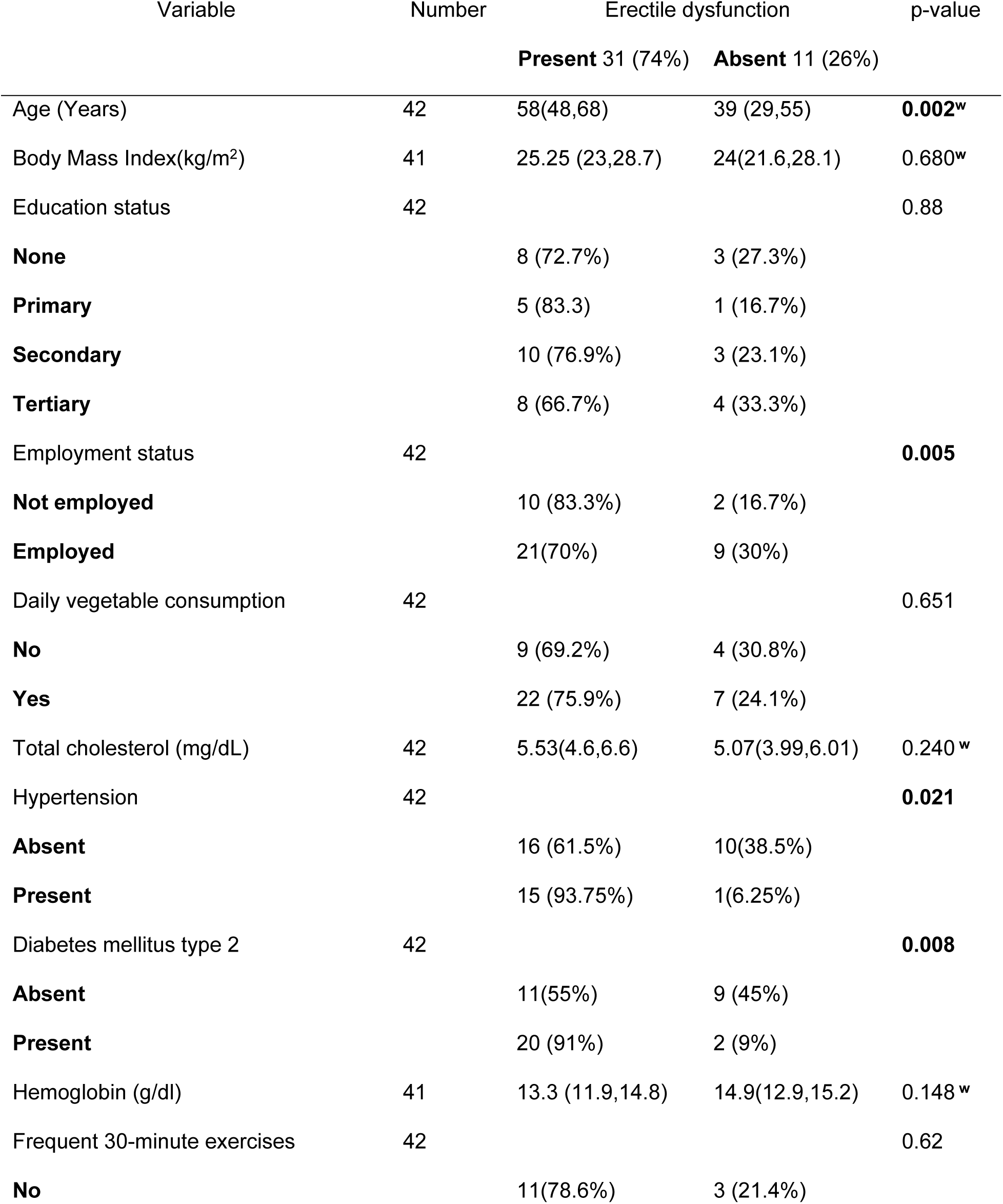
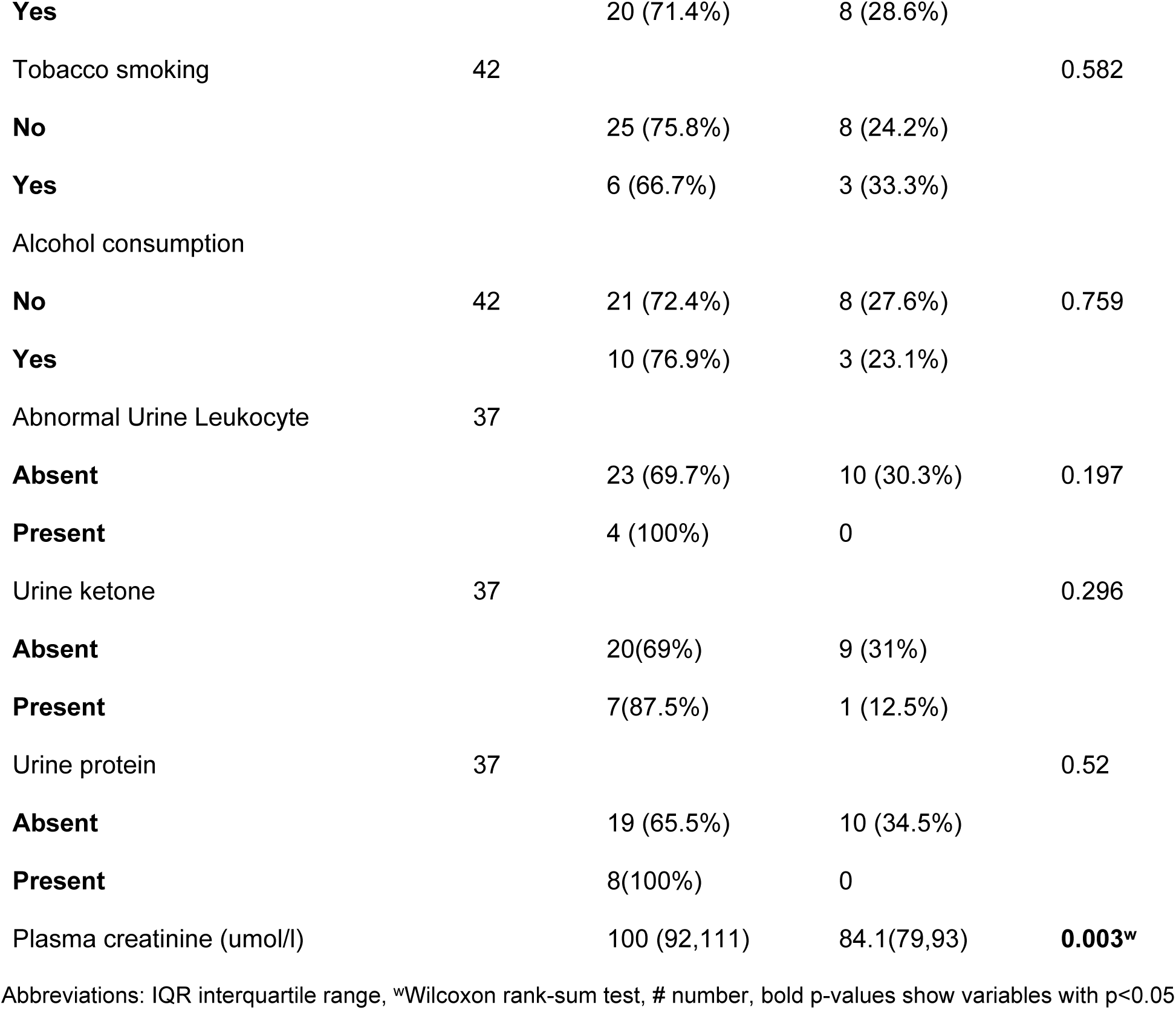
Relationship between ED and demographic and clinical characteristics.

### Univariable and multivariable analysis

Table 3 below shows the univariable and multivariable analyses investigating the factors associated with ED. At univariable analysis, a year increase was significantly associated with a 10% increased odds of experiencing ED, odds ratio (OR) 1.10, 95% CI 1.02 - 1.17, p = 0.005. Employed individuals had significantly lower odds of experiencing ED compared to their unemployed counterparts, OR 0.10, 95% CI 0.01 - 0.58, p = 0.010. The presence of hypertension and diabetes were also found to be significantly associated with increased odds of ED, both showing a strong association; OR of 9.37 and 8.18, respectively. Additionally, higher levels of plasma creatinine were associated with an increased risk of ED, OR 1.09, 95% CI 1.02 - 1.17, p = 0.010.

**Table 3:** Univariable and Multivariable analysis of factors associated with ED.

| Variable | Univariable analysis |  |  | Multivariable analysis |  |  |
| --- | --- | --- | --- | --- | --- | --- |
|  | OR | 95%CI | p-value | OR | 95%CI | p-value |
| **Age (years), median (IQR)** | 1.10 | 1.02 - 1.17 | **0.005** | 1.04 | 0.92 - 1.17 | 0.463 |
| **Employment status** |  |  |  |  |  |  |
| Unemployed | Ref |  |  |  |  |  |
| Employed | 0.10 | 0.01 - 0.58 | **0.010** | 0.01 | 0.00 - 0.84 | **0.041** |
| **Hypertension** |  |  |  |  |  |  |
| Absent | Ref |  |  |  |  |  |
| Present | 9.37 | 1.06 - 82.35 | **0.044** | 21.48 | 0.44 - 1046.94 | 0.122 |
| **Diabetes** |  |  |  |  |  |  |
| Absent | Ref |  |  |  |  |  |
| Present | 8.18 | 1.49 - 44.77 | **0.015** | 4.85 | 0.35 - 66.50 | 0.237 |
| **Plasma creatinine ($\mu\text{mol/l}$)** | 1.09 | 1.02 - 1.17 | **0.010** | 1.22 | 1.03 - 1.45 | **0.021** |

At multivariable analysis, employment status and plasma creatinine were significantly associated with ED. Employed individuals had lower odds of erectile dysfunction compared to the unemployed, OR 0.01, 95% CI 0.00 - 0.84, p = 0.041. A umol/l increase in plasma creatinine was significantly associated with a 3% increased chance of having erectile dysfunction, OR 1.22, 95% CI 1.03 - 1.45, p = 0.021.

## Discussion

The study aimed to investigate the prevalence and factors associated with ED in male patients with and without T2DM at LUTH.

The study reveals a high overall prevalence of ED (74%) among male participants. A substantial burden was among T2DM individuals (91%) compared to non-diabetic participants (55%), suggesting a strong connection between T2DM and ED. Several previous studies have also found ED to be higher in T2DM males compared to non-T2DM males ^10–12^. The higher prevalence of ED among T2DM participants emphasizes the need for targeted interventions within diabetes care. Clinically, healthcare providers should incorporate routine assessments for ED in their management of T2DM patients, considering it as an integral aspect of their overall health ^20–22^. Early identification of ED in this population can serve as a crucial marker for underlying cardiovascular and metabolic complications, prompting timely interventions to mitigate these risks ^23^. However, in our study, there was no statistical significance between T2DM and ED in multivariate analysis. It is worth noting that statistical significance or non-significance does not always denote biological significance. ^24,25^. Our study was underpowered in that it had a relatively low sample size (n=42) and as such the statistical significance could have been influenced by random error.

Employment status was significantly associated with ED. Unemployed individuals were more likely to experience ED compared to their employed counterparts. The association remained significant even after adjusting for other variables. This study finding was consistent with several previous studies which found that unemployed individuals were more likely to have ED ^26–28^. The finding implies that employment status may have a unique impact on ED risk, possibly related to stress, lifestyle, or other unexplored factors ^29,30^. Since ED has been shown by previous studies to be influenced by several factors including mental stress, psychogenic stress resulting from socioeconomic hardships may probably drive the development and progression of ED ^28,31^. The finding also highlights the need to incorporate psychosocial counseling in the management of ED in both T2DM and non-T2DM males.

Higher levels of plasma creatinine were consistently associated with an increased risk of ED in both univariable and multivariable analyses. Increased plasma creatinine is associated with kidney disease, which has been implicated in the development of ED ^32–35^. Chronic kidney disease is associated with progressive endothelial damage which compromises penile blood flow during arousal ^36^. Additionally, kidney disease is associated with hormonal imbalances that adversely affect the regulation of sexual function^35^. The combination of impaired vascular responses and hormonal irregularities amplifies the likelihood of ED in individuals with kidney disease ^37^. Previous studies have shown that T2DM males are at higher risk of developing chronic kidney disease which in turn is a risk factor for ED, a fact which partly explains the reason why T2DM males are at higher risk of having ED ^38^.

### Strengths and limitations

Our study demonstrates notable strengths that contribute to the robustness of its findings. Firstly, the use of a validated diagnostic tool, the International Index of Erectile Function (IIEF-5), stands out as a significant strength. This ensures a standardized and reliable assessment of ED, enhancing the accuracy and credibility of the study’s outcomes. The employment of such a well-established tool underscores the methodological rigor applied in diagnosing and measuring the phenomenon under investigation.

Secondly, the study serves as a pilot investigation, offering foundational data for future research endeavors. By establishing a baseline of information, particularly in the context of T2DM males, the study lays the groundwork for subsequent studies. This baseline not only aids in understanding the current landscape of ED in T2DM patients but also provides a platform for the development of improved healthcare strategies and interventions tailored to the unique needs of this population.

Despite these strengths, the study is not immune to certain limitations that warrant consideration. One prominent limitation is the relatively small sample size employed in the investigation. This size constraint raises concerns about the generalizability of the study’s findings to a broader population. Caution is therefore advised when attempting to extrapolate the results to diverse settings or populations, as the limited sample may not fully capture the variability present in the broader target group.

Another crucial limitation lies in the study’s reliance on self-reported information for several variables. Reliance on self-reported information introduces another layer of limitation. Key data, such as erectile function, alcohol consumption, and exercise habits, hinge on participants’ self-reports. This reliance on self-reporting may introduce biases, whether due to inaccuracies in recall or social desirability. Consequently, the precision and reliability of certain aspects of the study may be compromised, calling for a nuanced interpretation of the findings related to these self-reported variables.

## Conclusion

This study reveals a substantial burden of ED among the study participants, more especially in individuals with T2DM; and it was significantly associated with Plasma creatinine and employment status. This finding emphasizes the importance of comprehensive assessment and management of ED in T2DM patients. Further research with larger sample sizes and longitudinal designs is warranted to better understand the complex interplay of factors contributing to ED in this population and to develop targeted interventions for its prevention and management.

## Data Availability

All relevant data are within the manuscript and its Supporting Information files.

## Declarations

The authors declare no conflict of interest.

## Ethical considerations

Ethical approval was obtained from the Mulungushi University School of Medicine and Health Sciences Research Ethics Committee (SMHS-MU1-2023-24, dated 6th March 2023). Only participants who signed the consent were included in the study.

## Consent for publication

Not applicable

## Availability of data and materials

All data generated or analyzed in the study is included in this article. Other related data can be requested through the corresponding author.

## Competing interests

The authors declare that they have no competing interests

## Author’s contributions

SKM and LM conceived the study. LM, BCM, and BMH contributed to the writing of the manuscript. LM is the principal investigator. SKM is the senior author and supervisor. All authors read, provided feedback, and approved the final manuscript.

## Acknowledgments

We are grateful to the HAND research group at Mulungushi University School of Medicine and Health Sciences, the Laboratory staff, and the administration at Livingstone University Teaching Hospital for their strong support of the study.

## Notes

### Competing Interest Statement

The authors have declared no competing interest.

### Funding Statement

The author(s) received no specific funding for this work.

### Author Declarations

Ethical approval was obtained from the Mulungushi University School of Medicine and Health Sciences Research Ethics Committee (SMHS-MU1-2023-24, dated 6th March 2023).

